# A simulation-based approach for estimating the time-dependent reproduction number from temporally aggregated disease incidence time series data

**DOI:** 10.1101/2023.09.13.23295471

**Authors:** I Ogi-Gittins, WS Hart, J Song, RK Nash, J Polonsky, A Cori, EM Hill, RN Thompson

## Abstract

Tracking pathogen transmissibility during infectious disease outbreaks is essential for assessing the effectiveness of public health measures and planning future control strategies. A key measure of transmissibility is the time-dependent reproduction number, which has been estimated in real-time during outbreaks of a range of pathogens from disease incidence time series data. While commonly used approaches for estimating the time-dependent reproduction number can be reliable when disease incidence is recorded frequently, such incidence data are often aggregated temporally (for example, numbers of cases may be reported weekly rather than daily). As we show, commonly used methods for estimating transmissibility can be unreliable when the timescale of transmission is shorter than the timescale of data recording. To address this, here we develop a simulation-based approach involving Approximate Bayesian Computation for estimating the time-dependent reproduction number from temporally aggregated disease incidence time series data. We first use a simulated dataset representative of a situation in which daily disease incidence data are unavailable and only weekly summary values are reported, demonstrating that our method provides accurate estimates of the time-dependent reproduction number under those circumstances. We then apply our method to two previous outbreak datasets consisting of weekly influenza case numbers from 2019-20 and 2022-23 in Wales (in the United Kingdom). Our simple-to-use approach allows more accurate estimates of time-dependent reproduction numbers to be obtained during future infectious disease outbreaks.

## Introduction

An important challenge for policy makers during infectious disease outbreaks is to devise public health measures that limit transmission without placing an undue burden on the population [1–3]. Central to the decision making process is an ability to monitor changes in pathogen transmissibility in real-time during outbreaks, to determine whether current interventions are sufficient or whether additional restrictions are required.

A widely used measure of transmissibility is the time-dependent reproduction number (*R*_*t*_) [4–10]. The value of *R*_*t*_ represents the expected number of infections generated by someone infected at time *t* over the course of their entire infectious period. This quantity changes during an outbreak in response to interventions, variations in host behaviour and depletion of susceptible individuals due to infection-induced immunity. If the value of *R*_*t*_ is (and remains) below one, then the outbreak will decline. On the other hand, if the value of *R*_*t*_ is (and remains) above one, then the outbreak will grow.

Two distinct versions of *R*_*t*_ exist. First, the “instantaneous” reproduction number [4,5,11–13] represents the expected number of infections generated by someone infected at time *t*over their infectious period if transmission conditions do not change in future (i.e. assuming that the control interventions in place at time *t*, and any other factors that affect transmission, are not altered after time *t*). Second, the “case” reproduction number [12,14] is an analogous quantity but accounts for changes in transmissibility that occur after time *t*(due to, for example, changes in public health policy). Methods exist for estimating each of these versions of *R*_*t*_ [15]. However, here we focus on the instantaneous reproduction number as it is more amenable to analyses conducted in real-time during outbreaks when future changes in pathogen transmissibility are unlikely to be known. We therefore refer to the instantaneous reproduction number as *R*_*t*_ in this article.

A commonly used approach for estimating *R*_*t*_ is the method introduced by Cori *et al*. [4] (hereafter referred to as the Cori method), implemented in the R software package *EpiEstim* [16] and the online application *EpiEstim App* [17]. This approach is based on a renewal equation model of pathogen transmission (see Methods) and involves estimation of *R*_*t*_ from disease incidence time series and an estimate of the serial interval distribution (the probability distribution characterising the interval between symptom onset times in infector-infectee transmission pairs), both on a daily timescale. The Cori method has been extended in a range of ways following its original development [7], including accounting for imported cases [5,11,18,19], uncertainty in the serial interval distribution [5], superspreading [20,21], multiple pathogen variants [22] and unobserved generations of transmission [23].

However, a challenge that besets estimation of *R*_*t*_ using the Cori method is temporal aggregation of disease incidence time series data [7,24]. For COVID-19, for example, many public health agencies switched from publishing daily numbers of reported cases to weekly summaries after the height of the pandemic [25]. Often, disease incidence is reported weekly even for pathogens including influenza [26] for which realised serial intervals and generation times are typically only a few days [27–29]. A common workaround when using the Cori method in these scenarios is to match the timescale of the serial interval distribution to the timescale of the incidence data; for example, by supplying a weekly serial interval distribution and applying the Cori method to weekly incidence data. This is problematic not only because it is hard to unpick within-week changes in pathogen transmissibility when data are reported weekly, but also because an assumption of the transmission model underlying the Cori method is that all cases arising at timestep *t*are generated by infectors from earlier timesteps. In other words, if the Cori method is applied with a weekly timestep, as considered in this study, then it is assumed that an infector and infectee cannot both appear as cases in the disease incidence data in the same week. As the timescale of transmission (as characterised by the serial interval or generation time) of many pathogens is less than one week, this assumption is often incorrect when considering weekly aggregated disease incidence time series data.

In this research article, we address this issue by presenting a novel simulation-based method for estimating *R*_*t*_ from temporally aggregated disease incidence time series data and the serial interval distribution. Our approach involves repeated simulation of a renewal equation transmission model for different values of *R*_*t*_ with a timestep that is smaller than that of the disease incidence data. Using an iterative version of Approximate Bayesian Computation (ABC), we show how *R*_*t*_ can be estimated in real-time during outbreaks by matching model simulations exactly to the temporally aggregated outbreak data. We apply our approach to simulated data, demonstrating its accuracy and comparing results from our method to those obtained using the common workaround of the Cori method applied to temporally aggregated data. We go on to apply our method to real-world outbreak data from the 2019-20 and 2022-23 influenza seasons in Wales in the United Kingdom.

## Methods

In our analyses, we consider two possible approaches for estimating *R*_*t*_ from temporally aggregated disease incidence time series data: a workaround of the widely used Cori method (Approach 1 in Fig 1) and our novel simulation-based method (Approach 2 in Fig 1). Since the simulation-based approach uses a shorter timestep than that of data reporting, this method accounts for the possibility of multiple generations of transmission occurring between dates of data reporting. To provide a concrete setting in which to compare the two methods, we focus on a situation in which disease incidence data are aggregated into weekly timesteps.

**Figure 1.**
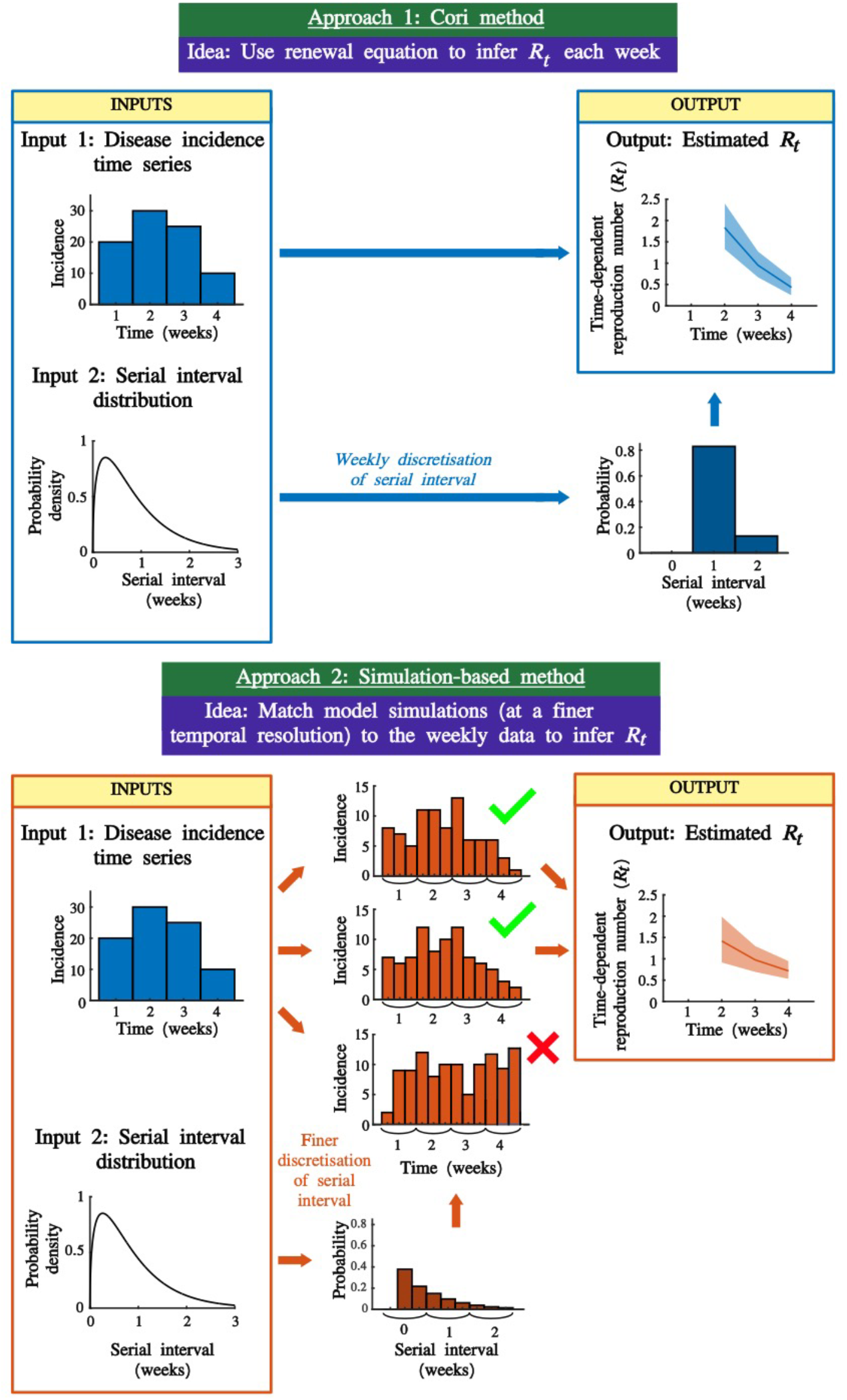
Schematic illustrating the approaches for estimating *R*_*t*_ that we consider. Approach 1 (top) involves the application of the commonly used Cori method to weekly aggregated disease incidence time series. Approach 2 (bottom) is the novel simulation-based approach, which involves matching simulations run with a smaller timestep to the weekly aggregated data to estimate *R*_*t*_. The second approach relaxes the assumption that individuals appearing in the incidence data cannot have infected other individuals appearing in the same week. Relaxing this assumption is particularly important during outbreaks in which the timescale of transmission is shorter than the temporal aggregation of the data (e.g. if disease incidence time series data are aggregated weekly, but serial intervals or generation times can be shorter than one week).

Below, we describe how the value of *R*_*t*_ each week can be estimated from the weekly data, first using the Cori method (with a timestep of one week, since the incidence data are aggregated into weekly values), and then using our simulation-based approach (using a timestep shorter than one week, again using the weekly incidence data).

### The Cori method

Following previous descriptions of the Cori method [4,5,11], we assume that the expected number of cases, *I*_*t*_, in week *t*, is given by

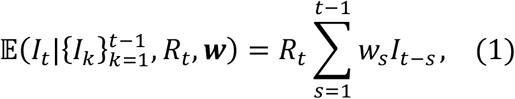

in which *w*_*s*_ is the probability that the (weekly discretised) serial interval takes the value *s* weeks. We use the notation ***w*** to denote the sequence of values of *w*_*s*_ (*s =* 1,2,…).

The goal of the Cori method is to estimate *R*_*t*_, assuming that it takes a constant value during the time period from week *t* − τ to week *t*. In our analyses, we set τ *=* 0 to obtain an estimate of *R*_*t*_ each week, but we first present the method for general (non-negative integer value) τ for consistency with previous presentations of this approach. If the number of cases in week *t*is drawn from a Poisson distribution, then the probability of observing weekly incidence 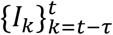 over the time window [*t* − τ, *t*] (which consists of incidence data from τ + 1 weeks) is

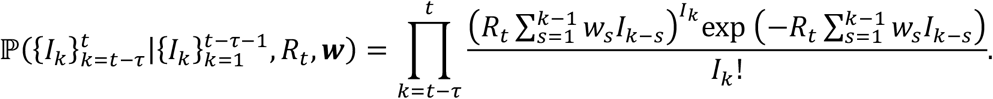

Assuming that the prior for *R*_*t*_ is a gamma distribution with shape parameter *α* and rate parameter *β*, then, by applying Bayes’ Theorem, the posterior for *R*_*t*_ is

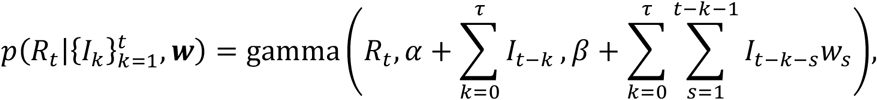

in which we use the notation 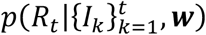 to represent the probability density function of *R*_*t*_ conditional on past incidence data and the weekly discretised serial interval distribution. The notation gamma(*x, a, b*) represents the probability density of a gamma distribution at value *x* with shape parameter *a* and rate parameter *b*. In all of our analyses, as in previous studies [4,5,11], we set *α =* 1 and *β =* 0.2. The prior for *R*_*t*_ therefore has mean and standard deviation equal to five. The large standard deviation is chosen so that the prior is relatively uninformative. The high mean ensures that the outbreak is not evaluated as being under control (*R*_*t*_ < 1) unless this is very likely to be the case, so that interventions are not relaxed erroneously.

Throughout the manuscript, we consider estimating individual values of *R*_*t*_ each week, based on the numbers of new cases observed in that week. In other words, as noted above, we assume that τ *=* 0, in which case the above expression simplifies to

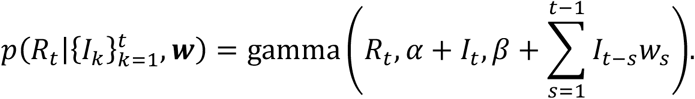

The Cori method can therefore be used to obtain a posterior for *R*_*t*_ for *t* ≥ 2 weeks.

### Simulation-based inference of *R*_*t*_

In the renewal equation model underlying the Cori method, the number of cases arising in week *t*depends on the numbers of cases in previous weeks. Implicit in that approach is an assumption that individuals appearing in the incidence data in any week cannot generate new cases in the same week. When disease incidence data are temporally aggregated, so that the timescale of transmission can be shorter than the timestep in the incidence data, this assumption may be incorrect. To relax this assumption, we consider a novel simulation-based approach for estimating *R*_*t*_. The goal of this method is again to estimate the value of *R*_*t*_ for each week, *t* ≥ 2, but using a renewal equation model with a timestep that is shorter than one week (e.g., a daily timestep).

#### Modified renewal equation

In this approach, we consider partitioning the cases in each week into *P* timesteps, where each new timestep is 1/*P* weeks. If, for example, *P =* 7, then we are using a daily timestep in the simulation-based method. We introduce the following notation:

- 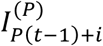 represents the number of cases in the *i*th (partitioned) timestep within week *t*(*i =* 1,2,…, *P*).
- 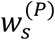 represents the probability that the serial interval, discretised into timesteps of length 1/*P* weeks (see below and Supplementary Material), takes the value *s* timesteps.
- ***w***^(***P***)^ represents the sequence of values of 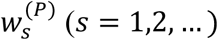.

In forward simulations of the corresponding renewal equation model, we assume that the number of cases in the *i* th timestep of week *t*is drawn from a Poisson distribution with mean

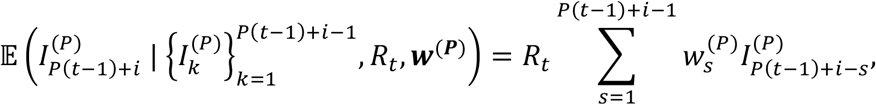

for *i =* 1,2,…, *P*. This is analogous to simulating the renewal equation underlying the Cori method but with a shorter timestep of 1/*P* weeks (rather than with a timestep of length one week).

#### Inference of R_t_

Inference of *R*_*t*_ under the simulation-based method involves repeated simulation of the modified renewal equation model, using an iterative version of ABC. In short, the model is simulated repeatedly in each week *t*, with a different value of *R*_*t*_ used in each simulation (these *R*_*t*_ values are sampled independently from the prior, and incidence data for times before week *t*are sampled from matching simulations from earlier weeks). This process is repeated until a fixed number of simulations (denoted *M*) have been run in which the simulated number of cases in week *t*exactly matches the corresponding number of cases in the data, *I*_*t*_. The values of *R*_*t*_ used to generate the matching simulations are then combined into a posterior estimate for *R*_*t*_. In all of our analyses using the simulation-based method, a value of *M =* 1000 was used.

This procedure is repeated iteratively, starting with *t=* 2, then *t=* 3, and so on. Since this approach only involves obtaining matching simulations for a single week at a time, estimates of *R*_*t*_ can be obtained relatively quickly (compared to attempting to match an entire simulation run over multiple weeks to the real-world data, as in standard ABC rejection sampling [30]). For a more detailed description of the simulation-based inference method, including an explanation of how cases are distributed between timesteps within the first week in each simulation, see the Supplementary Material. A schematic explaining the steps involved in the inference procedure is shown in Fig S1.

### Outbreak datasets

We consider three outbreak datasets in our analyses. We first test our approach on a simulated dataset. The use of simulated data not only enables us to compare estimates of *R*_*t*_ obtained using the simulation-based approach against analogous estimates using the Cori method, but it also allows us to verify that the simulation-based approach for estimating *R*_*t*_ generates accurate estimates in a setting in which we know true value of *R*_*t*_ (i.e., the value used to generate the simulated dataset). We then go on to compare outputs from the simulation-based approach and the Cori method using weekly aggregated disease incidence time series for influenza from 2019-20 and 2022-23 in Wales.

#### Simulated dataset (Fig 2)

We generated simulated data using the modified renewal equation, using a very small timestep so that the discretised serial interval is a close approximation to the continuous serial interval. Specifically, a disease incidence time series was generated starting from one initial case (in the first timestep) using a timestep of 10 minutes (*P =* 24 × 7 × 6 *=* 1,008). To generate a classic epidemic curve, the simulation was run for 11 weeks with *R*_*t*_ *=* 1.5 for *t* ≤ 6 weeks and *R*_*t*_ *=* 0.75 for *t* > 6 weeks.

**Figure 2.**
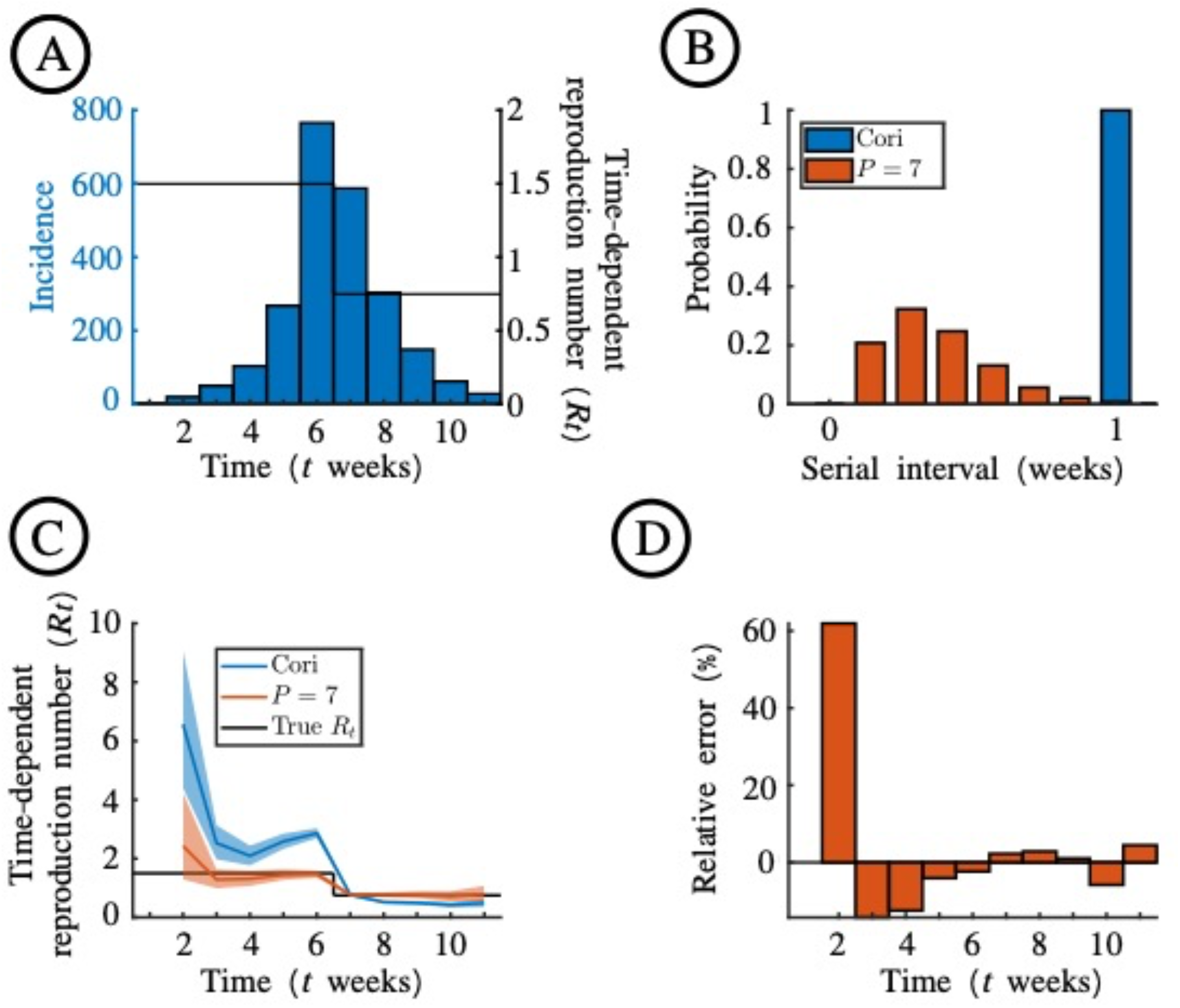
Estimation of *R*_*t*_ from the simulated disease incidence time series dataset. A. The simulated outbreak dataset (blue bars), generated with *R*_*t*_ *=* 1.5 for *t* ≤ 6 weeks and *R*_*t*_ *=* 0.75 for *t* > 6 weeks (black line). The outbreak was simulated with *P =* 1,008 starting from one initial case in the first timestep, and new cases were then aggregated into weekly case counts. B. The discretised serial interval, for *P =* 1 (as used with the Cori method; blue) and *P =* 7 (red). C. Estimates of *R*_”_ using the Cori method (blue) and the novel simulation-based approach (with *P =* 7; red). Blue and red lines are the mean estimates, and the shaded regions represent the 95% credible intervals. The value of *R*_*t*_ underlying the simulation is shown in black. D. The percentage error in the mean estimate of *R*_*t*_ each week (relative to the true value of *R*_”_ used to generate the dataset) using the simulation-based method with *P =* 7.

#### Influenza in Wales, 2019-20 (Figs 3,*4) and 2022-23 (Figs 5,6)*

To demonstrate our approach on real-world data, we considered two disease incidence time series datasets provided by Public Health Wales describing estimated numbers of cases of influenza-like illness (ILI) in Wales each week. The original data comprised the clinical consultation rate per 100,000 individuals in sentinel practices in Wales each week [31]. The total number of weekly cases was then estimated by multiplying each value in the original data by 31.075 (i.e. scaling these values based on the population size of Wales, which is 3,107,500 [32]). Since the dataset is for ILI, it likely contains some cases that were not influenza. Nonetheless, these data are sufficient to demonstrate and test the methods that we present in our study, and so we assume that the datasets are representative of numbers of influenza cases in Wales. Weekly data were provided from 28 October 2019 to 2 February 2020 (Fig 3A) and 31 October 2022 to 5 February 2023 (Fig 5A). These date ranges each span 14 weeks with high ILI burden.

**Figure 3.**
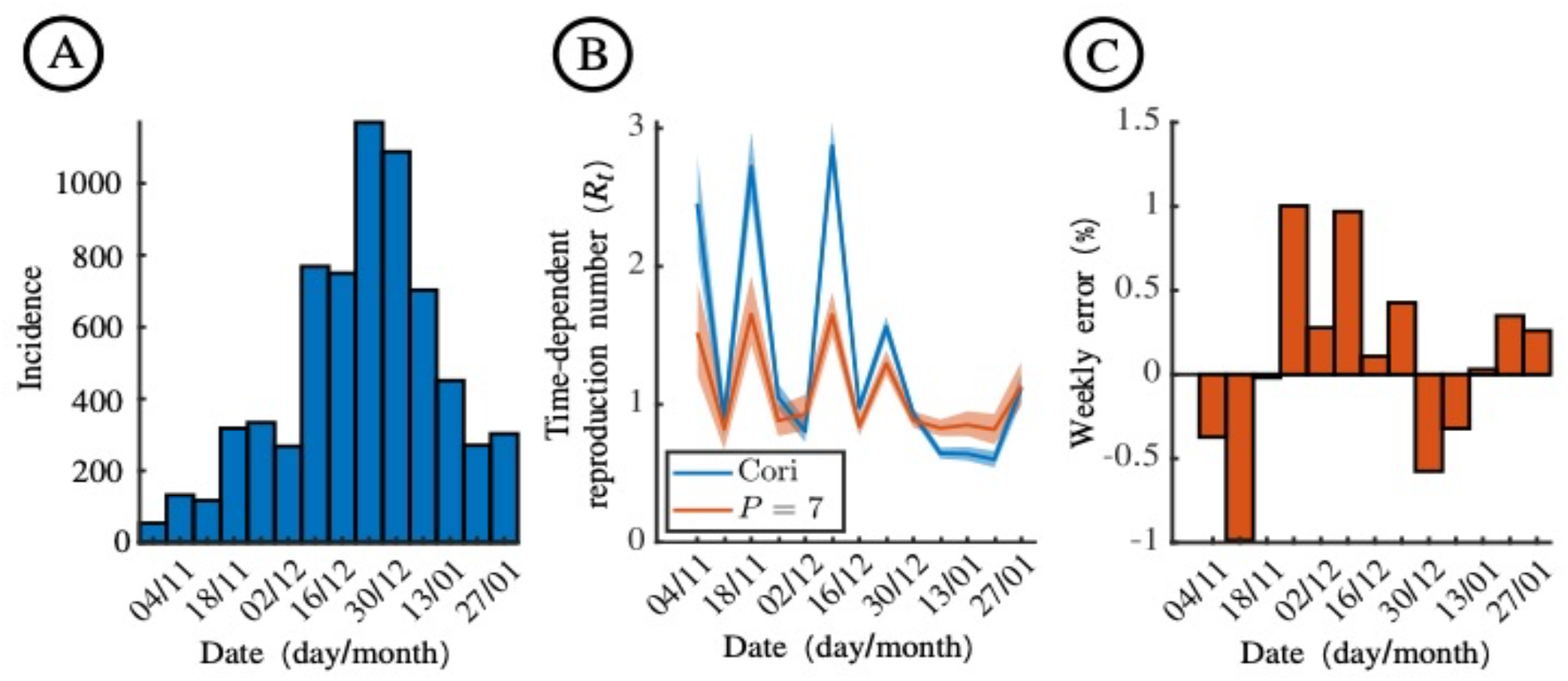
Estimation of *R*_*t*_ for influenza in Wales, 2019-2020. A. Weekly numbers of ILI cases in Wales from 28 October 2019 to 2 February 2020, estimated from surveillance data collected in sentinel practices. B. Estimates of *R*_*t*_ using the Cori method (blue) and the novel simulation-based approach (with *P =* 7; red). Blue and red lines are the mean estimates, and the shaded regions represent the 95% credible intervals. C. The percentage error in the mean estimate of *R*_*t*_ each week using the simulation-based method with *P =* 7, compared to using a larger value of *P =* 168 (which corresponds to estimating *R*_*t*_ with a one-hour timestep).

**Figure 4.**
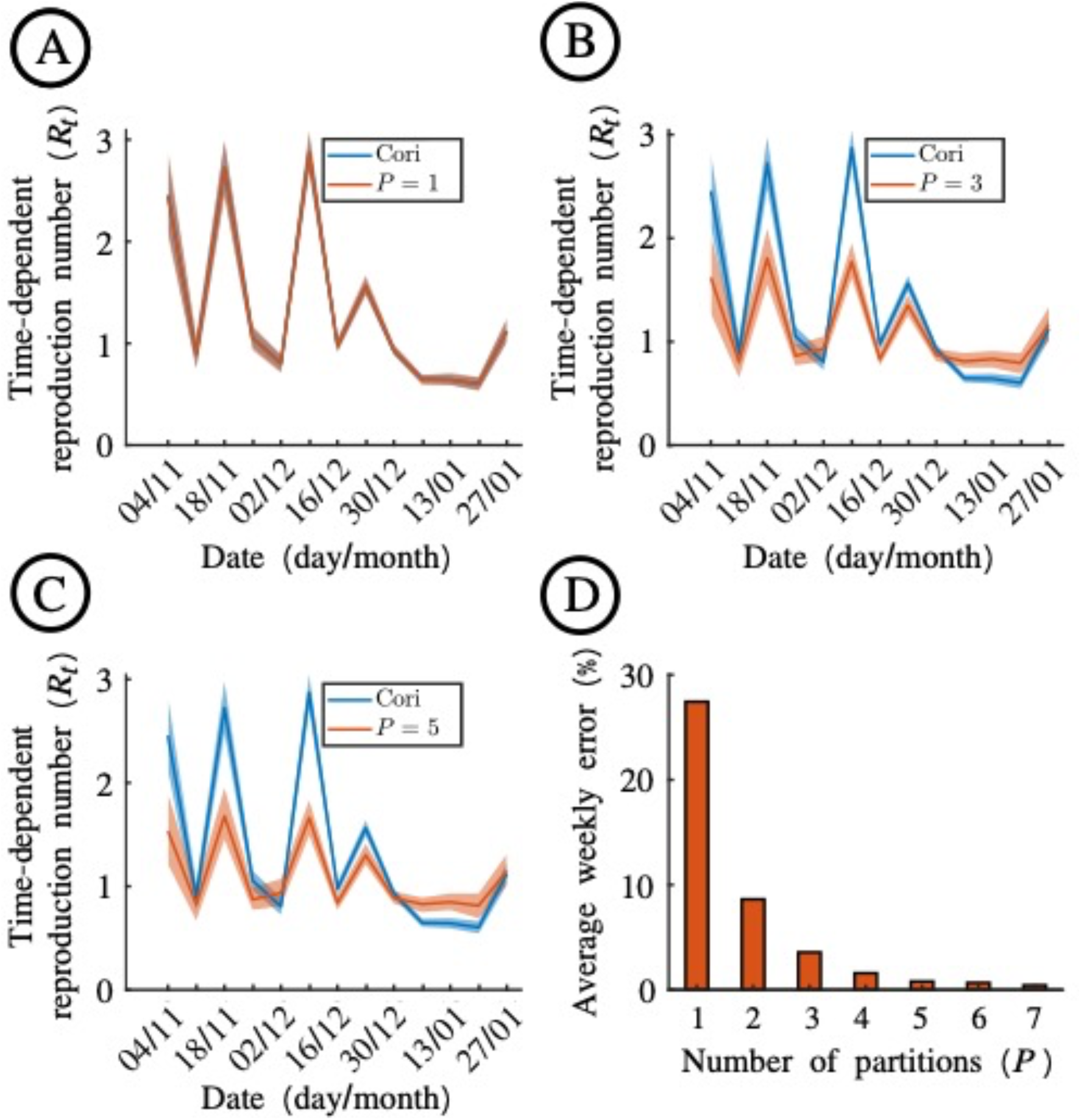
Dependence of *R*_*t*_ estimates using the simulation-based method on the value of *P* used, for influenza in Wales, 2019-2020. A. Estimates of *R*_*t*_ obtained when the Cori method (blue) and the novel simulation-based approach with *P =* 1 (red) are applied to the 2019-20 influenza dataset (Fig 3A). B. Analogous to panel A, but with *P =* 3 in the simulation-based approach. C. Analogous to panel A, but with *P =* 5 in the simulation-based approach. D. The average weekly absolute error in mean *R*_*t*_ estimates obtained using the simulation-based method with different values of *P*, compared to using a larger value of *P =* 168 (which corresponds to estimating *R*_*t*_ with a one-hour timestep). For a given value of *P*, this measure represents the absolute value of the error in the estimate of *R*_*t*_ in week *t*(compared to using *P =* 168), averaged over all values of *t*.

**Figure 5.**
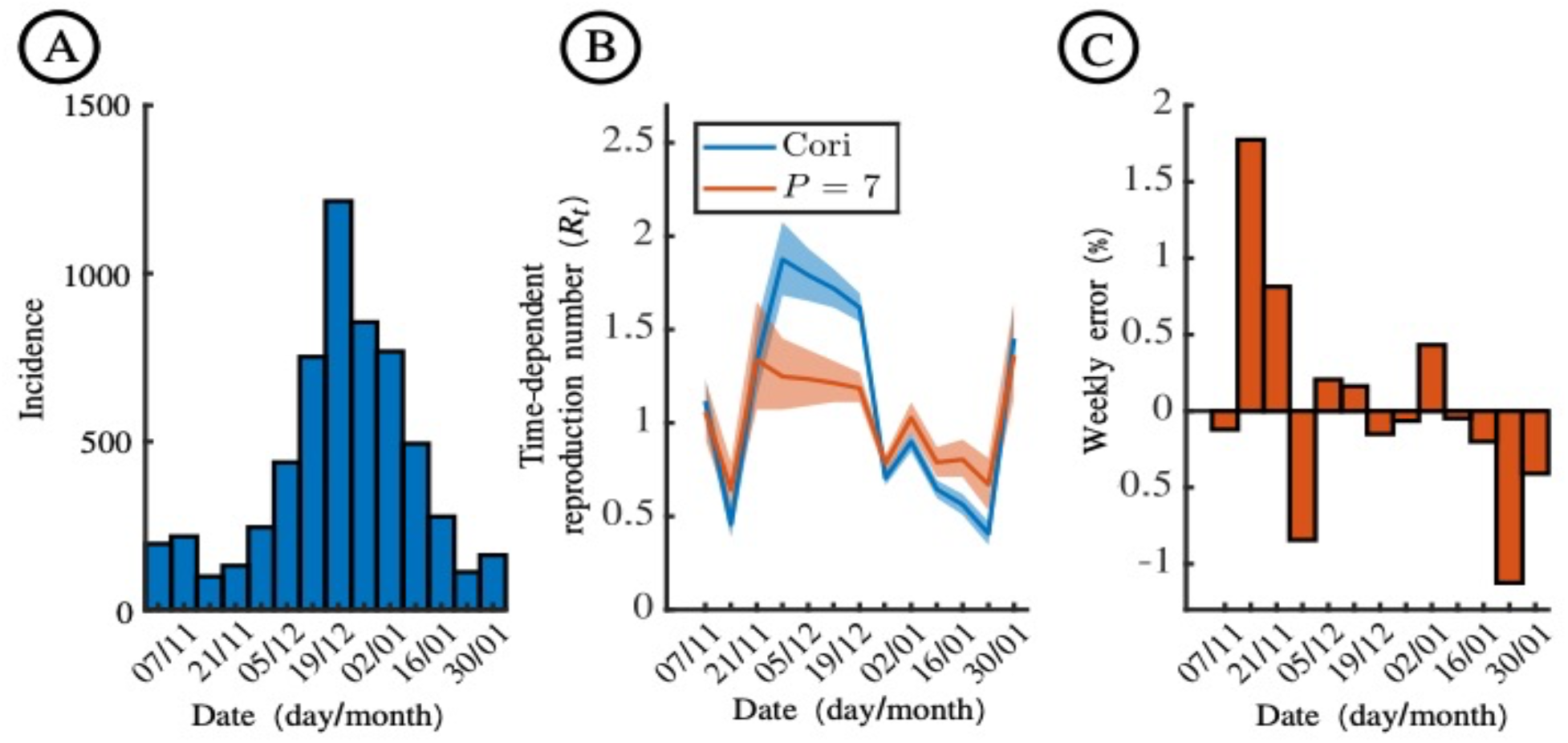
Estimation of *R*_*t*_ for influenza in Wales, 2022-2023. A. Weekly numbers of ILI cases in Wales from 31 October 2022 to 5 February 2023, estimated from surveillance data collected in sentinel practices. B. Estimates of *R*_*t*_ using the Cori method (blue) and the novel simulation-based approach (with *P =* 7; red). Blue and red lines are the mean estimates, and the shaded regions represent the 95% credible intervals. C. The percentage error in the mean estimate of *R*_”_ each week using the simulation-based method with *P =* 7, compared to using a larger value of *P =* 168 (which corresponds to estimating *R*_*t*_ with a one-hour timestep).

**Figure 6.**
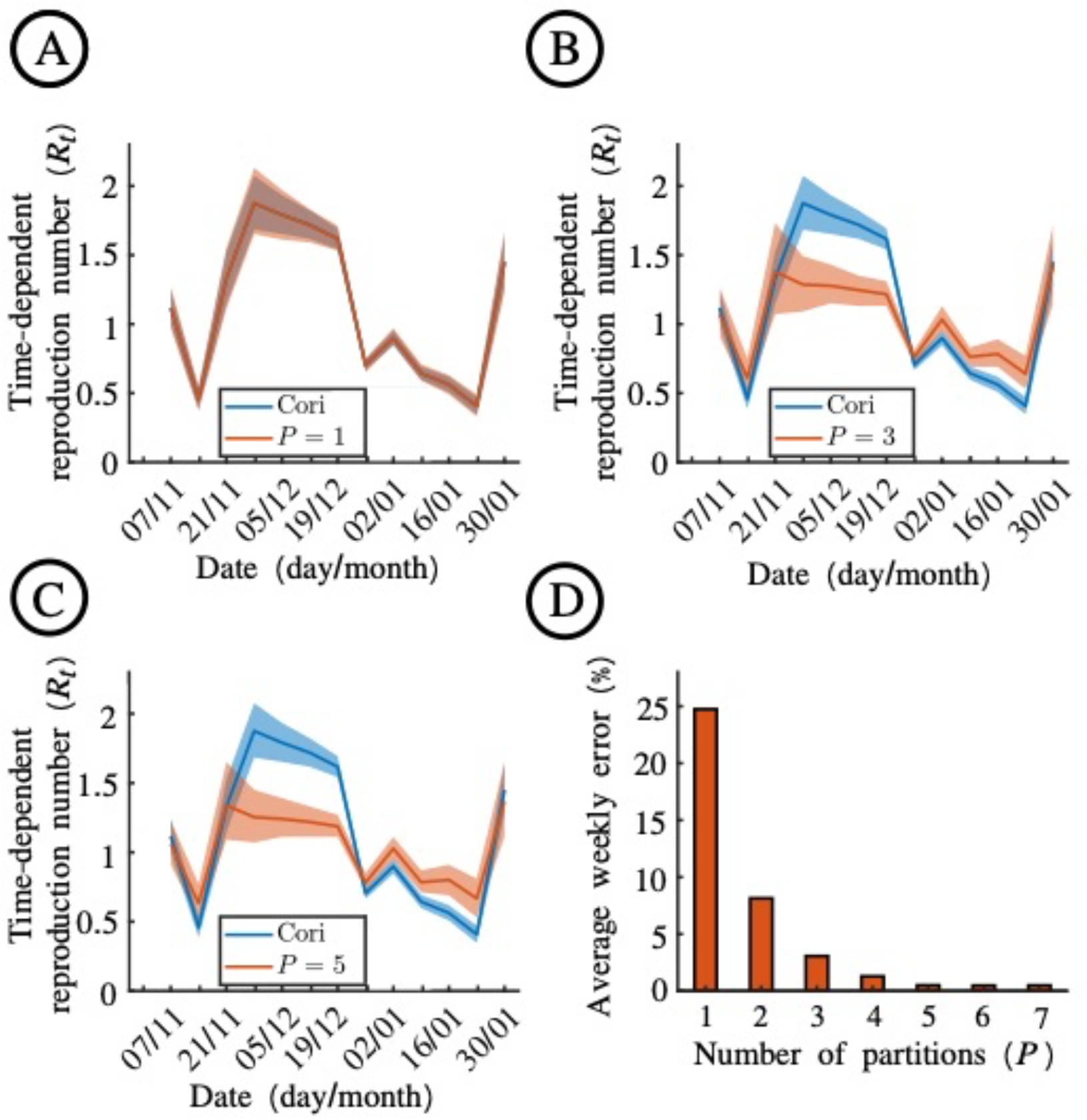
Dependence of *R*_*t*_ estimates using the simulation-based method on the value of *P* used, for influenza in Wales, 2022-2023. A. Estimates of *R*_*t*_ obtained when the Cori method (blue) and the novel simulation-based approach with *P =* 1 (red) are applied to the 2022-23 influenza dataset (Fig 5A) B. Analogous to panel A, but with *P =* 3 in the simulation-based approach. C. Analogous to panel A, but with *P =* 5 in the simulation-based approach. D. The average weekly absolute error in mean *R*_*t*_ estimates obtained using the simulation-based method with different values of *P*, compared to using a larger value of *P =* 168 (which corresponds to estimating *R*_”_ with a one-hour timestep). For a given value of *P*, this measure represents the absolute value of the error in the estimate of *R*_”_ in week *t*(compared to using *P =* 168), averaged over all values of *t*.

### Serial interval

Since we analyse influenza outbreak datasets in this study, we assume throughout that the (continuous) serial interval distribution is a gamma distribution with mean 0.37 weeks (2.6 days) and standard deviation 0.19 weeks (1.3 days) [33]. While this estimate was derived from household data for pandemic influenza, our focus is on demonstrating the application of the simulation-based method rather than precise estimation of the serial interval, and we expect this estimate to be in line with the serial interval for seasonal influenza (i.e., a mean value of less than one week). Denoting the probability density function of the serial interval distribution by *g*(*x*), then *g*(*x*) *=* gamma(*x*, 4, 10.8).

We discretise this distribution into timesteps of length 1/*P* weeks to obtain ***w***^(***P***)^. To do this, we adapt the method used by Cori *et al*. [4] in which the serial interval distribution is discretised into timesteps of length one. Specifically, we set

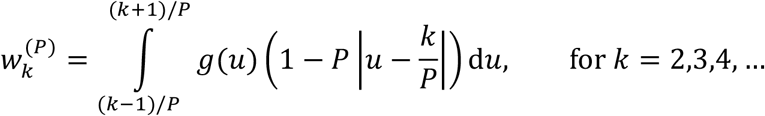

as derived in the Supplementary Material. We then choose 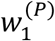 so that ***w***^(***P***)^ is a valid probability distribution (i.e., the sum of the entries of ***w***^(***P***)^ is one). The rationale for normalising ***w***^(***P***)^ in this way is that same-timestep cases (i.e. infectors and infectees appearing in the same timestep) are not possible in the renewal equation model. Our approach involves assigning all probability density near zero in *g*(*x*) to 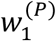, which is the shortest possible serial interval in the model.

## Results

### Simulated dataset

We first considered the simulated disease incidence time series dataset in which the incidence data are aggregated into weekly counts (Fig 2A). Since this dataset was generated using a serial interval for influenza, transmission occurred on a timescale less than one week. The discretised serial interval is shown in Fig 2B, both with a weekly timestep for use with the Cori method (*P =* 1; blue) and with a daily timestep for use with the simulation-based method (*P =* 7; red). Since the renewal equation model underlying both the Cori method and our simulation-based approach does not allow individuals appearing in the incidence data to generate new cases in the same timestep, only the simulation-based approach allows within-week realised serial intervals.

We applied both *R*_*t*_ inference methods to the simulated dataset, finding in this scenario that the simulation-based approach generates more accurate estimates of *R*_*t*_ than the Cori method (Fig 2C). The percentage error in the estimated value of *R*_*t*_ each week using the simulation-based approach with *P =* 7 (compared to value of *R*_!_ used to generate the dataset) is shown in Fig 2D.

In addition to our main analysis shown in Fig 2, we also conducted other analyses using the simulated dataset. We demonstrated that when the simulation-based method is applied with *P =* 1, the output matches the results obtained when the Cori method is used to estimate *R*_!_ (Fig S2A), as would be expected since the assumptions underlying the two methods are identical in this case. We also considered how *R*_!_ estimates obtained using the simulation-based method change when different values of *P* are chosen (Fig S2B-D), finding that the method can obtain accurate estimates for relatively small values of *P* (using a value of *P =* 3 led to similar errors compared to using *P =* 7).

### Influenza in Wales, 2019-20 and 2022-23

We then went on to consider the two Wales influenza outbreak datasets, again using both the Cori and simulation-based methods to estimate *R*_*t*_. First, we considered the weekly case counts from the 2019-20 influenza season (Fig 3A). As with the simulated dataset, the simulation-based approach led to different estimates of *R*_*t*_ than the Cori method; *R*_*t*_ estimates obtained using the simulation-based approach were typically lower than those from the Cori method during November and December 2019 (the simulation-based approach generally led to *R*_*t*_ estimates between one and two, whereas the Cori method generated estimates above two in multiple weeks), but then higher than those from the Cori method for most of January 2020 when *R*_*t*_ was estimated to be less that one (Fig 3B). We computed the percentage error in the *R*_*t*_ estimate each week using the simulation-based method with *P =* 7 (Fig 3C). Since the true underlying value of *R*_*t*_ was unknown, the percentage error was computed relative to applying the simulation-based method with a very large value of *P =* 168 (this is representative of the best possible estimate of *R*_*t*_ obtainable from the weekly incidence data; using a partitioning value of *P =* 168 returns inferred values of *R*_*t*_ estimated with a one-hour timestep). We also explored how *R*_*t*_ estimates depend on the value of *P* that is used (Fig 4). Estimates obtained using the Cori method and using the simulation-based method with *P =* 1 again matched closely (Fig 4A). We found that a value of *P =* 7 is large enough for accurate inference of *R*_*t*_ (Fig 4D).

The analyses of the Wales influenza data from 2019-20 were then repeated for the data from 2022-23, with similar results (Figs 5,6). Notably, the Cori method generally led to a higher estimate of *R*_*t*_ than the simulation-based method when *R*_*t*_ was estimated to be greater than one, and a lower estimate of *R*_*t*_ than the simulation-based method when *R*_*t*_ was estimated to be less than one (see Discussion).

## Discussion

During infectious disease outbreaks, evaluation of time-varying changes in pathogen transmission is essential to inform outbreak responses. Different metrics can be tracked, including incidence of new cases, hospitalisations and deaths, and outbreak growth rates [34,35]. A key metric that has been estimated in real-time during outbreaks of a range of pathogens is *R*_*t*_ in part because of its straightforward interpretation [7,9,15]. Not only is there a threshold value of *R*_*t*_ *=* 1, below which an outbreak can be inferred as being under control, but the value of *R*_*t*_ also provides information about the extent to which the level of transmission must change (relative to current transmission) for an outbreak to grow or decline. For example, if *R*_*t*_ *=* 2, then more than half of transmissions must be prevented for the outbreak to decline. Similarly, if *R*_*t*_ *=* 0.5, then up to twice as many transmissions may occur before the outbreak begins to grow. Precise estimation of *R*_*t*_ is therefore crucial.

Here, we have presented a novel simulation-based approach for estimating *R*_*t*_ in scenarios in which disease incidence time series data are aggregated temporally (Fig 1). While epidemiological data may be collected at a fine temporal resolution, it is common for the data to then be aggregated (e.g., into weekly or monthly counts). Aggregated data may be easier to report and can be more accurate than data presented at a high temporal resolution when there is uncertainty in the precise times at which cases occurred. However, as we have shown, frequently used methods for inferring *R*_*t*_, such as the Cori method [4,5], may not generate accurate estimates when applied to temporally aggregated data if transmission occurs more rapidly than the temporal resolution of the aggregated data. This is because the renewal equation model underlying the Cori method involves assuming that an individual appearing in the disease incidence time series data at timestep *t*cannot have infected other individuals appearing in the same timestep. Our proposed simulation-based approach addresses this, by exactly matching simulations of a renewal equation model run with a shorter timestep (*P* timesteps for each timestep in the aggregated data) to the temporally aggregated incidence data. The simulation-based approach not only provides accurate estimates of *R*_*t*_ (Fig 2), but can also be applied easily to real-world datasets (Figs 3-6). While using a very large value of *P* allows the most accurate possible *R*_*t*_ estimates to be obtained from the aggregated data, even relatively small values of *P* are sufficient for *R*_*t*_ to be inferred accurately (Figs 4,6).

We found that, while the Cori method did not always provide an accurate estimate of *R*_*t*_ due to the temporal aggregation of the disease incidence data, it was able to identify whether or not *R*_*t*_ is below one (i.e., the outbreak is under control). While this is useful, as noted above precise estimation of *R*_*t*_ is important as it provides information about the number of transmissions that must be prevented for an outbreak to be controlled (or the number of transmissions that can occur for an outbreak to remain under control). The difference in *R*_*t*_ estimates between the two methods can be explained by the assumption of no same-timestep cases (i.e., infectors and infectees cannot appear in the disease incidence time series in the same timestep) in the renewal equation. When the Cori method is applied to weekly data, this then leads to overestimation of the serial interval, which is known in turn to lead to overestimation of *R*_*t*_ if the true value of *R*_*t*_ is greater than one and underestimation of *R*_*t*_ if the true value of *R*_*t*_ is less than one [36,37].

A closely related study by Nash *et al*. [24], undertaken at the same time as the analyses presented here, has also considered estimation of *R*_*t*_ from temporally aggregated disease incidence time series data. In that approach, an expectation-maximisation (EM) algorithm is used to reconstruct daily incidence from any aggregation of disease incidence data using the serial interval (on a daily timescale). The original version of the Cori method is then applied to the estimated daily data. This EM approach has been integrated into the EpiEstim R software package [16]. There are several differences between the approach by Nash *et al*. [24] and the simulation-based method described here. First, the two approaches are methodologically distinct, relying on entirely different underlying methods (EM or model simulation). Second, under the approach by Nash *et al*., only a single estimated daily disease incidence time series is obtained. In contrast, our method involves matching a range of simulations to the temporally aggregated data, thereby considering different possible disaggregated disease incidence time series that could have led to the weekly aggregated data. Third, our method can be run straightforwardly for a range of values of *P*, allowing the most accurate possible estimates of *R*_*t*_ to be inferred from temporally aggregated incidence data (discretisation into a timestep of less than one day is straightforward). Fourth, our approach can be applied easily in scenarios in which the serial interval distribution is discrete rather than continuous (if, for example, the serial interval distribution is constructed directly from observations of dates on which infector-infectee pairs report symptoms). A rigorous comparison of estimated values of *R*_*t*_ obtained using the simulation-based method and using the EM approach of Nash *et al*. [24] is a target for future exploration. However, an initial investigation involving applying the methods to the Wales influenza datasets considered here suggests that the two approaches can obtain consistent results (Fig S3).

Our simulation-based method is conceptually straightforward, simply requiring repeated simulation of a renewal equation model. It is also computationally efficient to run, as simulations are only required to match the real-world data for one aggregated timestep at a time. This contrasts with using ABC rejection sampling to estimate all values of *R*_*t*_ simultaneously, which would involve matching entire simulated time series to the entire real-world dataset. The efficiency of our approach allowed us to require that the simulations used to infer *R*_*t*_ match the real-world data exactly. Further computational efficiency could be achieved by removing this condition, and instead setting a threshold “distance” within which a simulation is determined to match the real-world data, as is common when using ABC [30,38]. However, this necessitates that a distance metric is chosen, and resulting estimates of *R*_*t*_ may be less accurate.

As in any modelling study, our framework in its current form involves assumptions. We followed previous publications in which the Cori method has been used [4,5] and assumed that the time series datasets from which we estimated *R*_*t*_ represent numbers of new symptomatic cases in each timestep. In the disease incidence time series data, it is then assumed that each infectee appears after their infector following a time period that reflects a random draw from the serial interval distribution, which is assumed to always take strictly positive values. However, in reality, realised serial intervals can be negative (if an infectee develops symptoms before their infector; this is possible, for example, for transmission of SARS-CoV-2 [39–42]). Rather than using disease incidence time series, it is possible to apply both the Cori method and the simulation-based method to data describing incidence of infections, replacing the serial interval distribution as an input with the distribution of the generation time (the interval between infection times in infector-infectee pairs). This can be beneficial as realised generation times are always positive. However, since times of infection are often unknown, infection incidence data are typically not observed directly. Consequently, further inference would then be required to estimate incidence of infections, as well as to estimate the generation time distribution [8,43–45].

In our analyses, we assumed that all cases in the disease incidence time series (after the first timestep) arose because of transmission within the population under consideration, and that all cases were recorded. In reality, some infected individuals may become infected outside the local population [5,11,46,47] and under-reporting of cases is likely for many pathogens [48–51]. Extension of our method to account for these features of real-world outbreaks is a target for future research. Similarly, our method assumes that a Poisson distributed number of cases occur in each timestep of the modified renewal equation model. Considering different possible probability distributions, including accounting for the possibility of superspreading events on some days [20,21,52], is another possible area for future work.

Further testing of the performance of the simulation-based approach in different scenarios would also be worthwhile. For example, in settings in which disease incidence time series are subject to a “day-of-the-week effect” [53] (e.g., if cases occurring at the weekend are typically reported with a longer reporting delay than those arising during the week), *R*_*t*_ inference using the simulation-based method applied to weekly aggregated incidence data may generate more robust estimates than attempting to infer *R*_!_ from less accurate daily incidence data. Our method can also be adapted for scenarios in which the disease incidence time series data are aggregated into timesteps that are not all of equal length. For example, when incidence data are derived from World Health Organization reports that are published irregularly in time, the timestep changes during the outbreak [54], and those irregular timesteps can be used directly in our simulation-based method.

In summary, we have presented a novel method for estimating *R*_!_ from temporally aggregated disease incidence time series. Going forwards, the ideal scenario is for disease incidence time series to be recorded accurately at a fine temporal resolution (e.g., daily). If that occurs, then existing methods for estimating *R*_*t*_ are generally expected to perform well. However, if disease incidence time series continue to be aggregated temporally for pathogens for which transmission occurs on a short timescale, then methods allowing accurate *R*_*t*_ inference from temporally aggregated data are of paramount importance.

## AUTHORS’ CONTRIBUTIONS

IOG – formal analysis, investigation, visualisation, validation, writing – original draft, writing – review and editing.

WSH – methodology, writing – review and editing.

JS – methodology, writing – review and editing.

RKN – methodology, writing – review and editing.

JP – methodology, writing – review and editing.

AC – methodology, writing – review and editing.

EMH – methodology, supervision, visualisation, writing – review and editing.

RNT – conceptualization, methodology, project administration, supervision, visualisation, writing – original draft, writing – review and editing.

## ACKNOWLEDGEMENTS

Thanks to the Communicable Disease Surveillance Centre at Public Health Wales for providing us with the data used in this research. Thanks to members of the Zeeman Institute for Systems Biology and Infectious Disease Epidemiology Research at the University of Warwick, particularly Alex Kaye, and the Wolfson Centre for Mathematical Biology at the University of Oxford, for useful discussions about this work.

## DATA AVAILABILITY

The computing code used to perform the analyses in this article is available in the following GitHub repository: www.github.com/billigitt/R_Estim_Simulation_Method. All computer code was written in the MATLAB programming environment (compatible with version R2022a).

