## Supplementary Material for "A simulation-based approach for estimating the time-dependent reproduction number from temporally aggregated disease incidence time series data"

### Supplementary Text

#### Discretisation of the serial interval

Here, we explain how a continuous serial interval distribution (with probability density function $g(x)$) can be discretised into timesteps of length $1/P$ weeks to obtain $w_s^{(P)}$ ($s = 1,2,\ldots$) and $\boldsymbol{w}^{(\boldsymbol{P})}$. The notation $w_s^{(P)}$ represents the probability that the serial interval, discretised into timesteps of length $1/P$, takes the value $s$ timesteps, and $\boldsymbol{w}^{(\boldsymbol{P})}$ is the sequence of values of $w_s^{(P)}$. We adapt the approach described by Cori *et al.* [1] (see web appendix 11 in the Supplementary Data of that article) in which the serial interval is discretised into timesteps of length one.

We consider an infector-infectee transmission pair and assume that the precise time at which the infector develops symptoms is uniformly distributed within the timestep in which they appear in the disease incidence time series data. If the continuous serial interval takes the value $u$ weeks, then the probability that the infectee arises in the disease incidence time series data $k \geq 2$ timesteps after their infector is given by

$$\mathbb{P}(\text{discrete SI} = k \mid \text{cts SI} = u) = \begin{cases} 1 - P\left|u - \frac{k}{P}\right|, & \text{if } \frac{k-1}{P} < u < \frac{k+1}{P}, \\ 0, & \text{otherwise.} \end{cases}$$

Then, conditioning on the unknown value of the continuous serial interval gives

$$w_k{}^{(P)} = \int_0^\infty \mathbb{P}(\text{discrete SI} = k \mid \text{cts SI} = u) \times g(u)\mathrm{d}u,$$

$$= \int_{(k-1)/P}^{(k+1)/P} \left(1 - P\left|u - \frac{k}{P}\right|\right) g(u)\,\mathrm{d}u,$$

in which $g(u)$ is the probability density function of the continuous serial interval distribution.

In principle, the calculation above can be applied when $k = 1$, and a similar argument can be used to obtain the probability that an infectee appears in the disease incidence time series in the same timestep as their infector (which would correspond to $w_0{}^{(P)}$). However, since the renewal equation model requires all new cases in a given timestep to have been infected by infectors appearing in the incidence data at a strictly earlier timestep, rather than the same timestep, we neglect $w_0{}^{(P)}$ and instead assume that same timestep cases are absorbed into $w_1{}^{(P)}$. In other words, we simply set $w_1{}^{(P)}$ so that $\boldsymbol{w}^{(\boldsymbol{P})}$ sums to one.

When we apply the Cori method, we require the continuous serial interval distribution to be discretised into weekly timesteps. This therefore corresponds to undertaking the above calculations with $P = 1$.

**Simulation-based inference of $R_t$**

Here, we give further details about the simulation-based method. The value of $R_t$ (for $t \geq 2$) is estimated iteratively: in other words, $R_2$ is estimated first, followed by $R_3$, and so on. By estimating $R_t$ iteratively, our inference procedure can be performed more quickly than attempting to estimate $R_t$ for all values of $t \geq 2$ simultaneously (as in standard ABC rejection sampling [2]).

To estimate $R_t$ (for $t \geq 2$) from a weekly disease incidence time series dataset, we consider running simulations of the modified renewal equation model in which each week is divided into $P$ timesteps (each of timestep $1/P$ weeks). The value $P = 7$ therefore corresponds to a daily timestep, however the simulation-based method can be run for any positive integer value of $P$ (with larger values of $P$ leading to the most accurate possible estimates of $R_t$ obtainable from the weekly aggregated disease incidence time series).

To estimate $R_2$, we repeatedly simulate the modified renewal equation up until the end of the second week, storing "matching" simulations (those simulations in which the number of cases in the second week in the simulation exactly matches the number of cases in the second week in the time series dataset). In each simulation, we: i) sample the value of $R_2$ from the (time-homogeneous) prior for $R_t$; ii) assign each case in the first week of the dataset to one of the $P$ timesteps in the first week (chosen uniformly at random). New simulations are generated until $M$ simulations that match the number of cases in the second week of the dataset have been obtained. For each matching simulation, we store both the sampled value of $R_2$ and the corresponding numbers of cases in each timestep in that simulation, $\{I_i^{(P)}\}_{i=1}^{2P}$. The values of $R_2$ from the matching simulations can be combined to construct the posterior distribution for $R_2$.

We then estimate $R_t$ for each $t \geq 3$ in turn. To do this, we again run simulations of the modified renewal equation model, but starting from the beginning of week $t$ (this corresponds to timestep $P(t-1)+1$ in the modified renewal equation model). Each simulation is run until the end of week $t$ (i.e. up to and including timestep $Pt$). In each simulation, we: i) sample the value of $R_t$ from the prior; ii) choose past incidence uniformly at random out of the matching sets stored when estimating $R_{t-1}$. New simulations are generated until $M$ simulations that match the number of cases in week $t$ of the dataset have been obtained. For each matching simulation, we store both the sampled value of $R_t$ and the corresponding numbers of cases in each timestep in that simulation (including the sampled past disease incidence used in that simulation), $\{I_i^{(P)}\}_{i=1}^{Pt}$. The values of $R_t$ from the matching simulations can be combined to construct the posterior distribution for $R_t$.

In all of our analyses, we required simulations that match the disease incidence time series data in week $t$ to have exactly the correct number of cases in that week. For improved computational efficiency, this algorithm could be adapted so that the number of cases in week $t$ in matching simulations is within some tolerance level of the corresponding number of cases in the real-world data. However, we did not use that approach here as it would lead to less accurate estimates of $R_t$, and we found that our computing code ran sufficiently quickly for results to be obtained without this adaptation.

### Supplementary Figures

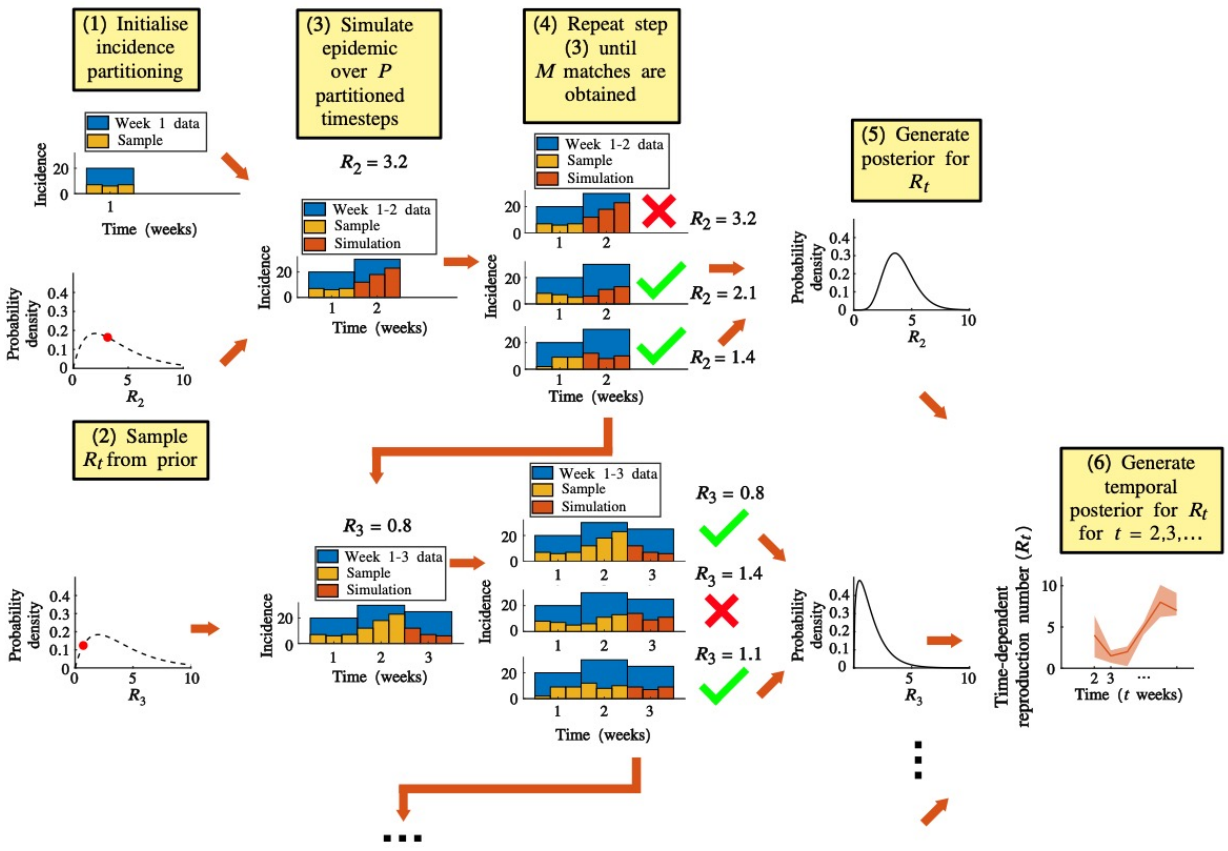


**Fig S1. Schematic illustrating the steps involved in the simulation-based method for inferring $R_t$.** The procedure involves six steps: (1) Initialise the incidence partitioning for the first aggregated timestep ($t = 1$). Each case in the first aggregated timestep is assigned uniformly at random to one of the $P$ partitions in that aggregated timestep; (2) Sample the value of $R_2$ from the prior; (3) Use the partitioned incidence from step 1 and the $R_2$ value from step 2 to simulate the partitioned incidence in week $t = 2$ using the modified renewal equation. (4) Repeat steps 1-3 until a pre-specified number of simulations, $M$, have been generated in which the simulated number of cases in week $t = 2$ match the corresponding number of cases in the disease incidence data. (5) Use the sampled values of $R_2$ from the matching simulations to construct the posterior distribution for $R_2$. These steps are then repeated iteratively to estimate $R_t$ in each of weeks $t = 3,4,5$ ... For these values of $t$, in each simulation, the value of $R_t$ is sampled from the prior, and step 1 is replaced so that past disease incidence for times up to (and including) week $t-1$ are sampled from the matching simulations obtained when estimating $R_{t-1}$. (6) Plot the posterior distributions for $R_t$ ($t = 2,3,4$ ...) to observe temporal changes in transmissibility during the outbreak.

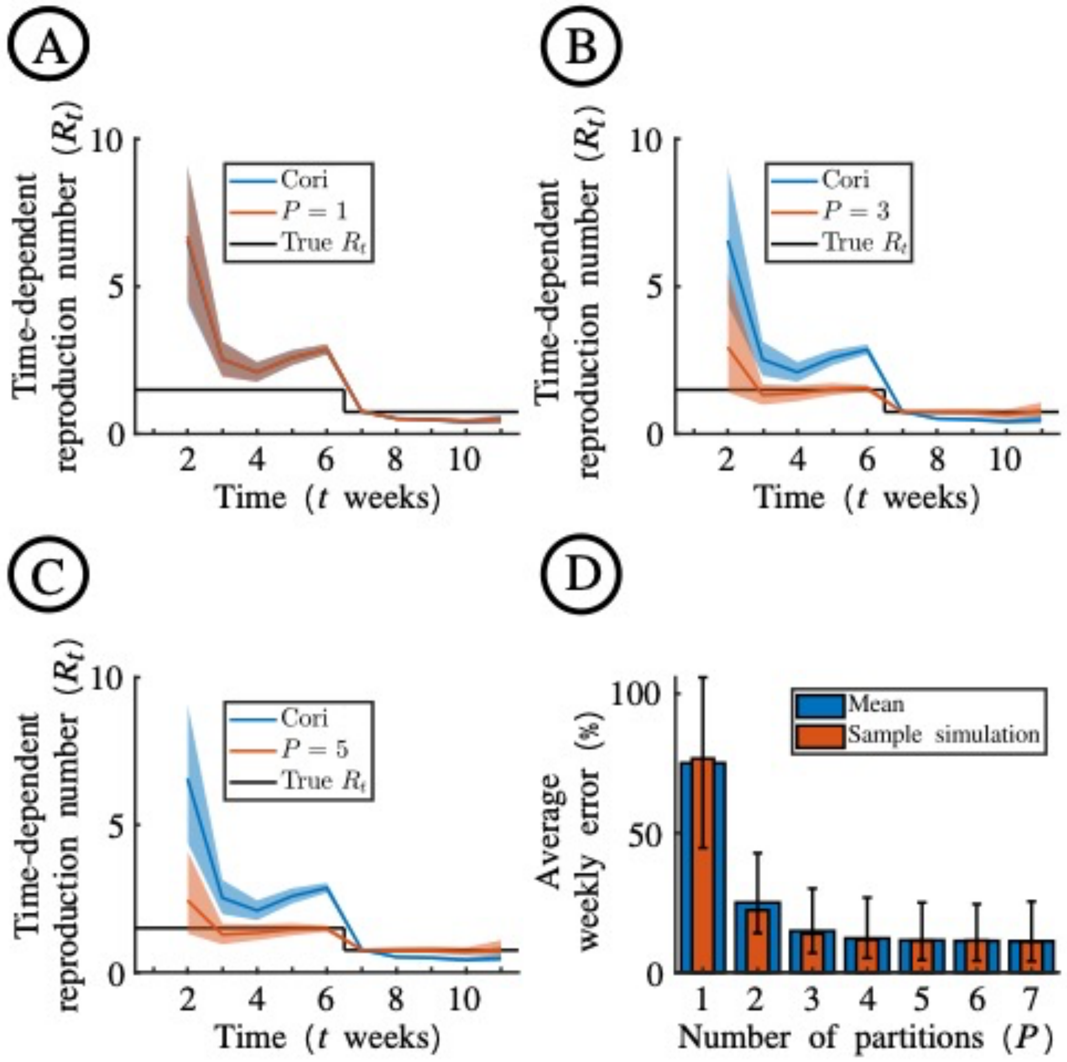


**Fig S2. Dependence of $R_t$ estimates using the simulation-based method on the value of $P$ used, for the simulated disease incidence time series dataset.** A. Estimates of $R_t$ obtained when the Cori method (blue) and the novel simulation-based approach with $P = 1$ (red) are applied to the simulated disease incidence time series dataset (Fig 2A in the main text). B. Analogous to panel A, but with $P = 3$ in the simulation-based approach. C. Analogous to panel A, but with $P = 5$ in the simulation-based approach. D. The average weekly absolute error in mean $R_t$ estimates obtained using the simulation-based method with different values of $P$, compared to the true underlying value of $R_t$. For a given value of $P$, this measure represents the absolute value of the error in the estimate of $R_t$ in week $t$ (compared to the true value of $R_t$), averaged over all values of $t$. Red bars are for the simulated dataset shown in Fig 2A of the main text. Blue bars are the average weekly absolute error averaged over each of 100 simulated datasets that were generated in an identical fashion to the simulated dataset in Fig 2A of the main text. Error bars show the 95% credible interval across the 100 simulations.

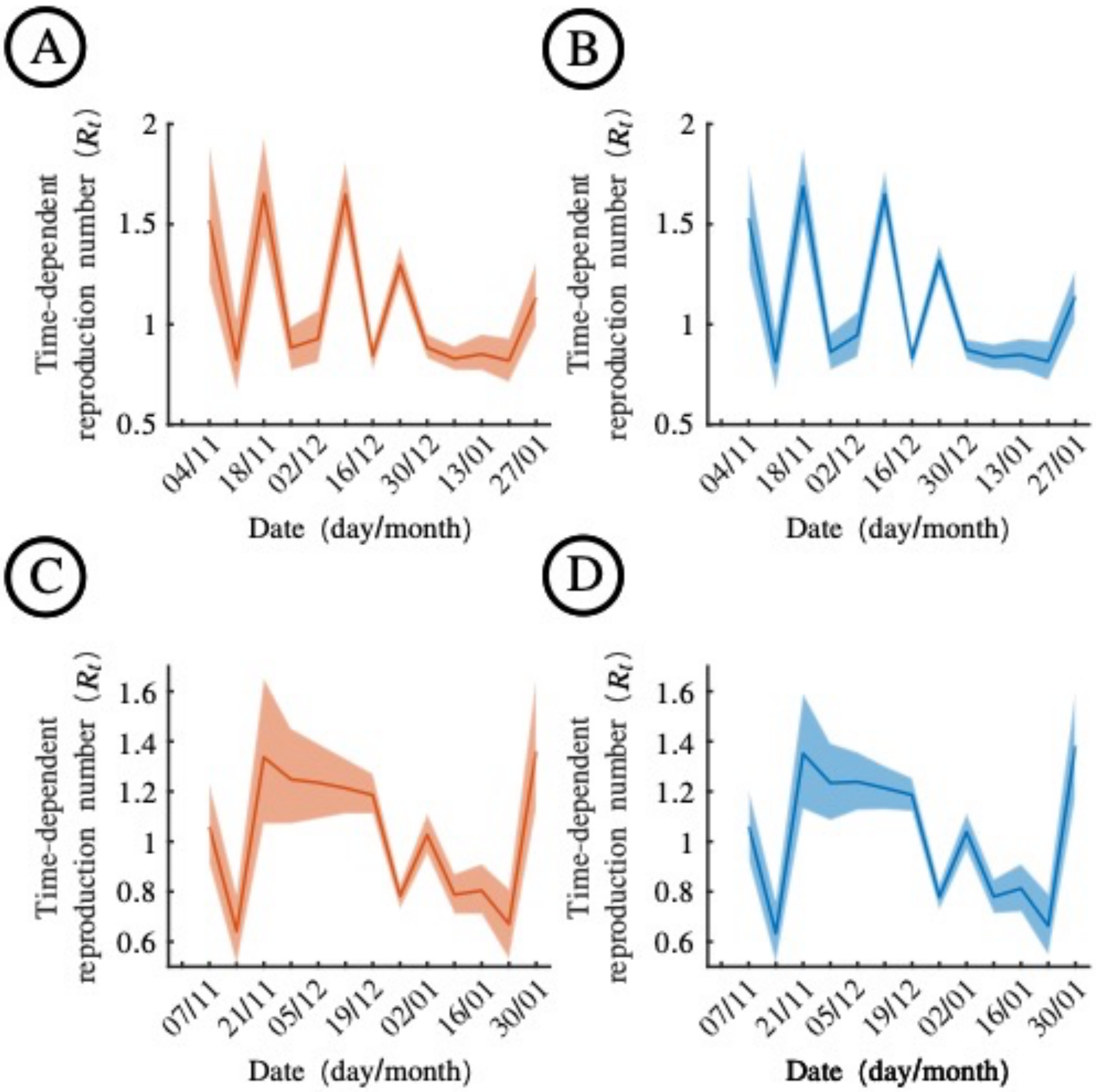


**Fig S3. Comparison of $R_t$ estimates obtained using our simulation-based approach with analogous estimates using the Expectation-Maximisation (EM) approach developed by Nash *et al.* [3].** A. Estimates of $R_t$ obtained when the simulation-based approach with $P = 7$ is applied to the 2019-20 Wales influenza dataset (Fig 3A). B. Analogous results to panel A, but using the EM approach. C. Estimates of $R_t$ obtained when the simulation-based approach with $P = 7$ is applied to the 2022-23 Wales influenza dataset (Fig 5A). B. Analogous results to panel C, but using the EM approach. Blue and red lines are the mean estimates, and the shaded regions represent 95% credible intervals. These results indicate that the simulation-based and EM approaches generate consistent results.
